# ddPCR Reveals SARS-CoV-2 Variants in Florida Wastewater

**DOI:** 10.1101/2021.04.08.21255119

**Authors:** Eben Gering, Jacob Colbert, Sarah Schmedes, George Duncan, Joe Lopez, Jessy Motes, James Weiss, Taj Azarian, Omer Tekin, Jason Blanton

## Abstract

Wastewater was screened for the presence of functionally significant mutations in SARS-CoV-2 associated with emerging variants of concern (VOC) by ddPCR, and results accorded with sequencing of clinical samples from the same region. We propose that PCR-based screening of wastewater can provide a powerful tool for rapid and inexpensive screening of large population segments for VOC-associated mutations and can hone complementary sampling and sequencing of direct (human) test material to track emerging VOC.

---

Several SARS-CoV-2 variants of concern have been identified that show elevated transmissibility (Tegally et al. 2020, Tu et al. 2021, Volz et al. 2021, Galloway et al. 2021, Lauring and Hodcraft 2021). The mechanisms driving this transmissibility are not fully understood, but select VOC-associated mutations have been found to strengthen viral binding to host cell receptors (Starr et al. 2020), inhibit antibody binding (Gupta et al. 2020) and increase viral replication within human tissues (Plante et al. 2020). Research is actively underway to assess whether newly emerging variants are linked to higher disease severity in infected humans and/or resistant to vaccine-induced immunity (Lauring and Hodcroft 2021, de Cosio et al. 2021). Regardless, characterizing the emergence and spread of VOC is crucial to pandemic mitigation because an increase in transmissibility means an increase in cases, and a heightened potential for further emergences of VOC. These effects may impact the success of vaccination programs designed to achieve herd immunity and end the COVID-19 pandemic. Intercontinental exchanges of VOC, and convergent evolution of VOC-associated mutations (Kemp et al. 2020, Gupta et al. 2021, Tu et al. 2021, Washington et al. 2021), further suggest that new variants will emerge within many communities before SARS-CoV-2 can be extirpated.

Investigators have recently deployed analyses of wastewater to overcome logistical and budgetary constraints posed by standard phylodynamic surveys of circulating SARS-CoV-2 genotypes (i.e., sequencing of individual patient samples). This approach leverages the discovery that SARS-CoV-2 infected hosts shed viral RNA in feces; this shedding also precedes symptom onset in those who develop COVID-19. Sewage samples have been used since the early stages of the ongoing pandemic to track prevalence trends and forecast impending cases (e.g., Bivens et al. 2020, Gonzalez et al. 2020, Peccia et al. 2020, Stadler et al. 2020). Furthermore, Jahn et al. (2021) recently deployed next-generation sequencing (NGS) of wastewater samples to identify SARS-CoV-2 VOCs in archival wastewater material, discovering that a highly transmissible emerging lineage (B.1.1.7) was established in Switzerland prior to its discovery within a patient sample. This work illustrates the broad utility of wastewater testing for VOC tracking, but NGS genomic analyses are also costly, time consuming, sensitive to inhibition, and challenging to interpret. More rapid and robust approaches to VOC screening of wastewater samples are urgently needed in order to enhance these sequencing efforts and leverage limited resources.

In the present study we utilize outsourced digital droplet PCR (ddPCR) analyses of wastewater to ascertain the emergence of two VOC-associated mutations in the RNA encoding SARS-CoV-2’s spike protein (N501Y and 69-70del) within raw wastewater samples from several wastewater facilities in South Florida. This approach employs a rapid assessment protocol for the identification of viral genotypes circulating in over a million residents, with ∼24-hour turnaround at extremely low cost. An additional advantage of this approach is that wastewater analyses provide egalitarian indexes of focal communities, circumventing problematic biases in direct testing (e.g., effects of an individual’s age, mobility, work status, Internet access, etc. on the likelihood of undergoing a standard COVID-19 test).

We targeted two mutations, N501Y and 69-70del, for screening in influent wastewater samples in January 2021. The first mutation, N501Y, appears to enhance SARS-CoV-2 transmission based on its rapid spread in SARS-CoV-2 lineages that emerged in parallel within the UK (B.1.1.7/501Y.V1) and South Africa (B.1.351/501Y.V2). N501Y is also found in an emerging variant first reported from Brazil (B.1.1.248/501Y.V3/P1) that was found in a Minnesota patient who was sampled on January 9, 2021, following travel to Brazil. N501Y involves a single replacement in the ACE2 binding motif that is predicted to increase the strength of binding between spike and hACE2-expressing host cells (Starr et al. 2020). We also screened wastewater samples for 69-70del, a 6bp deletion that occurs on several backgrounds including B.1.1.7. B.1.1.7 currently occurs at increasing frequencies across the United States (US). The first US case was reported on December 29, 2020, and a case within central Florida was reported just two days later. Washington et al. (2021) provided phylogenetic evidence that B.1.1.7 reached the US via multiple introductions starting at least as early as November 2020. The 69-70del that is found in B.1.1.7 is also found in other SARS-CoV-2 lineages, including two variants that show no indications of elevated transmissibility in humans: the Danish ‘mink strain’ and B1.375, the latter of which has been reported in a patchy distribution across the US via sequencing of patient swabs. Thus, of the two mutations we screened by ddPCR, N501Y is of highest concern since it involves functional changes expected to enhance hACE2 binding, whereas 69-70del is often, but not always, found in rapidly spreading VOC including B.1.1.7 and 501Y.V2.

**Figure 1.**
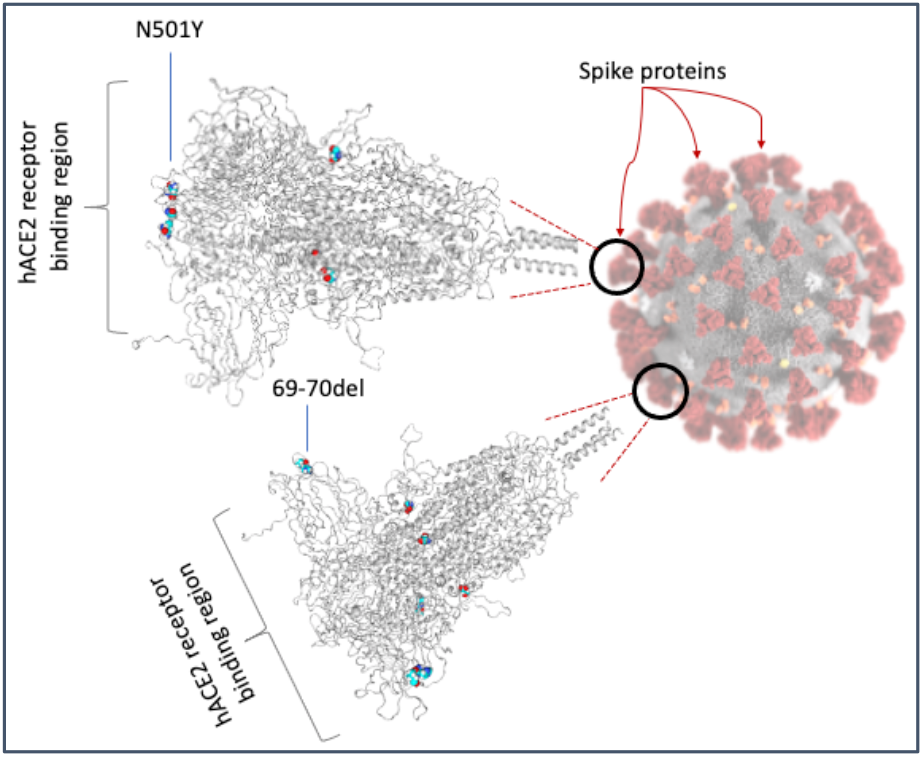
SARS-CoV-2 loci assayed by ddPCR in the present study rendered by COV3D (Gowthaman et al. *in press*). N501Y involves the spike protein’s receptor binding motif and is reported to: a) increase the strength of spike binding to human host cell receptors (hACE2) and b) inhibit the binding of select neutralizing host antibodies. N501Y occurs in multiple SARS-CoV-2 lineages within North America. 69-70del involves the deletion of two amino acids the spike protein. Like N501Y, 69-70del has evolved in multiple SARS-CoV-2 lineages including B.1.1.7 – a variant that contains several additional derived mutations inclusive of N501Y. The other color-filled mutation in the illustrations above is D614G which is now found at high frequency worldwide and associated with enhanced viral replication in respiratory tissues (Plante et al. 2020, Hou et al. 2020).

Wastewater-testing by ddPCR provided an inexpensive and rapid method of screening for the presence of VOC-associated mutations in large populations totaling over a million residents. Both mutations we screened for were identified at all four surveyed treatment locations (Table One). This demonstrates the widespread presence of emerging SARS-CoV-2 genotypes within the study region. The result was not surprising given that the US Center for Disease Control announced confirmed cases of B.1.1.7 (which contains both focal mutations) in California, central Florida, Colorado, Georgia, and New York on January 6, 2021, the same week our wastewater samples were collected. Additionally, a convergently-evolved strain containing the N501Y mutation was discovered in the US in December 2020 (Tu et al. 2020); infections involving this lineage and/or undiscovered clade(s) may underlie our findings of N501Y and 69-70del in regional wastewater.

**Table One:** Recovery-adjusted concentrations of SARS-CoV-2 in wastewater measured using ddPCR targeting the conserved (EUA) N1 locus and the proportion of droplets containing two focal mutations (N501Y and 69-70del) that are associated with emerging SARS-CoV-2 variants.

|  | Population serviced (N residents) | Recovery-adjusted SC2 (copies/L) | BCoV Recovery (%) | %N501Y | %69-70del |
| --- | --- | --- | --- | --- | --- |
| Source#1 | 631,000 | 946,673 | 5.88% | 4.926 | 1.370 |
| Source#2 | 338,000 | 2,187,749 | 7.17% | 9.122 | 2.471 |
| Source#3 | 226,000 | 1,148,302 | 4.95% | 3.284 | 1.943 |
| Source#4 | 131,000 | 2,190,138 | 7.35% | 2.279 | 0.529 |
| **Total population surveyed = 1,326,000** |  | **Weighted average N501Y = 5.03%**<br>**Weighted average 69-70del = 1.43%**<br>(weighting based on recovery-adjusted SC2 levels) |  |  |  |

At the present time, both N501Y and 69-70del are present at low frequency at each of the sampled localities. This pattern is suggestive of a generally low prevalence of infections involving VOCs upstream of the sampled sewage plants and comports with sequencing results from recent human clinical specimens. Florida Bureau of Public Health Laboratories sequenced 220 SARS-CoV-2-positive clinical samples from the same geographic region as the wastewater facilities from March 2020 through January 2021 as part of routine surveillance of SARS-CoV-2. N501Y was detected in one surveillance sample (0.45% frequency), and S69-70del was detected in five surveillance samples (2.27% frequency), which includes one sample from the B.1.1.7 lineage with both mutations. Five additional B.1.1.7 samples were detected from the same region during targeted screening for VOC in January 2021. It is too soon to tell if these variants are increasing over time in the study region; thus, continued monitoring and sequencing of both environmental and human samples should be a key research priority. We also note that shedding rates may differ between hosts who are infected with divergent SARS-CoV-2 genotypes. Until this has been established, frequencies of VOC and/or specific mutations found in wastewater (e.g. via ddPCR or sequencing) cannot provide precise estimates of the proportion of COVID-19 cases in a community that involve a mutation or lineage of interest.

Our wastewater analyses alone cannot determine which viral lineage(s) are present (and contain the two mutations detected via ddPCR), though clearly they include lineages other than B.1.1.7, given the unbalanced frequencies of N501Y and 69-70del in our samples. Identifying specific clades in North American wastewater samples will require RNA sequencing approaches (as in Jahn et al. 2021), and/or development of strain-specific ddPCR protocols. However, we have shown here that ddPCR assays of VOC markers can play a crucial role in focusing this work - pinpointing geographical regions or timepoints over which to prioritize SARS-CoV-2 sequencing. Looking ahead, we expect that a two-step approach involving PCR screening to identify sequencing targets at various scales, and subsequent genotyping of environmental or human samples will provide a highly efficient approach to phylodynamic epidemiology. This could be facilitated by nascent wastewater surveillance networks (e.g. those under development in the US and EU), open source protocols to screen for VOC-associated mutations (e.g. Vogels et al. 2021) and thriving research collaborations emerging worldwide, in parallel with VOC.

## Methods

24-hour flow-metered composite samples of primary influent wastewater were collected under refrigeration using autosamplers. Samples were held at 4°C and processed via extraction and ddPCR within ∼72-hour windows by a commercial laboratory (GT Molecular, Fort Collins Colorado). In brief, BCoV surrogate virus was added to each wastewater sample, and RNA was then isolated from 40mL aliquots for amplification of CDC’s EUA N1 locus target. Measured total SARS-CoV-2 concentrations (using the N1 locus) were then adjusted for effects of each wastewater matrix using recovery fractions determined via ddPCR amplification of the BCoV surrogate spike-in. Each sample was then subjected to ddPCR screening for the frequencies of the two targeted (VOC-associated) mutations.

### RNA from clinical specimens

Primary specimens were received at the Bureau of Public Health Laboratories, Florida Department of Health and confirmed for the presence of SARS-CoV-2 RNA. RNA was purified by multiple methods, including: Kingfisher combined with the Thermo Fisher MagMAX Viral/Pathogen II (MVP II) Nucleic Acid Isolation kit (Applied Biosystems by Thermo Fisher Scientific, Waltham, MA); Abbott *m*2000 System combined with the Abbott RealTi*m*e SARS-CoV-2 assay (Abbott, Lake County, IL;, MagNA Pure 96 combined with the MagNA Pure 96 DNA and Viral NA Small Volume Kit (Roche, Indianapolis, IN); MagNA Pure LC 2.0 combined with the MagNA Pure LC Total Nucleic Acid Isolation Kit (Roche, Indianapolis, IN); and EZ1 Advanced combined with the EZ1 DSP Virus Kit (Qiagen, Germantown, MD), QIAamp DSP Viral RNA Mini Kit (Qiagen, Germantown, MD).

### cDNA and amplicon generation

cDNA was synthesized from RNA with the following reaction: 10 µL template RNA, 6 µL nuclease-free water, 4 µL SuperScript IV VILO Master Mix (Thermo Fisher Scientific, Waltham, MA). cDNA synthesis reactions were incubated at: 25°C for 10 minutes, followed by 50°C for 10 minutes, and 85°C for 5 minutes. cDNA was amplified using each of the two ARTIC v3 primer pools (https://artic.network/ncov-2019) (Integrated DNA Technologies, Inc, Coralville, IA) which tile the SARS-CoV-2 genome. That reaction contained: 12.5 µL Q5 2x Master Mix (New England Biolabs, Ipswich, MA), 1 µL Primer pool (#1 or #2) (20 µM), 9 µL Nuclease-free water, and 2.5 µL cDNA. The amplicon reactions were incubated at: 98°C for 30 seconds, followed by 95°C for 15 seconds, and 64°C for 5 minutes (the last two steps were repeated a total of 35 times). Amplicon pools 1 and 2 were then combined, cleaned up with 1:1 Axygen^™^ AxyPrep Mag^™^ PCR Clean-up beads (Axygen, Union City, CA), and quantified by Qubit Fluorometer and Broad Range DNA assay (Thermo Fisher Scientific, Waltham, MA).

### Library generation and sequencing

Libraries were prepared from the amplicons using Illumina’s Nextera XT library prep kit (Illumina, San Diego, CA) and quantified by Qubit Fluorometer and Broad Range DNA assay and TapeStation capillary electrophoresis (Agilent, Santa Clara, CA). Equimolar amounts of each library were pooled and concentrated to 20 µL using 0.7x Axygen^™^ AxyPrep Mag^™^ PCR Clean-up beads. The final pooled sample was quantified using a Qubit Fluorometer and High Sensitivity DNA assay. To confirm the expected library size, pooled libraries were run on an Agilent TapeStation. Libraries were sequenced on either an Illumina MiSeq or NextSeq 550 Dx. For the MiSeq: The sample pools were diluted to 2 nM based on the Qubit measurements and Agilent sizing information, and 10 µL of the 2 nM pool was denatured with 10 µL of 0.2 N NaOH. Amplicon libraries were diluted to 8 pM in Illumina’s HT1 buffer, spiked with 5% PhiX, and sequenced using a MiSeq 600 cycle v3 kit. For the NextSeq 550Dx: The sample pools were diluted to 4 nM based on the Qubit measurements and Agilent sizing information, and 5 µL of the 4 nM pool was denatured with 5 µL of 0.2 N NaOH. Amplicon libraries were diluted to 1.5 pM in Illumina’s HT1 buffer, spiked with 1% PhiX, and sequenced using a NextSeq 500/550 Mid Output Kit v2.5 (300 cycle).

### SARS-CoV-2 genome assembly and variant detection

SARS-CoV-2 consensus assemblies were generated using a custom pipeline, FLAQ-SC2. Quality metrics were generated for raw sequence reads using fastqc v0.11.9 (http://www.bioinformatics.babraham.ac.uk/projects/fastqc/), and raw fastqs were quality filtered and trimmed (SLIDINGWINDOW:4:30 MINLEN:75 TRAILING:20) using trimmomatic v0.39 (Bolger, et al. 2014). Sequence adapter and Phix contamination were removed using bbduk (BBMap v38.79) (https://sourceforge.net/projects/bbmap/). Cleaned reads were mapped to SARS-CoV-2 reference NC_045512.2 using bwa mem v0.7.17-r1188.(Li, H. 2013). PCR duplicates were marked and removed using samtools v1.9.(Li, et al. 2009)). ARTIC primer sequences (https://github.com/artic-network/artic-ncov2019/tree/master/primer_schemes/nCoV-2019/V3) were trimmed from bam files using ivar trim v1.2.2 (Grubaugh, et al. 2019). Variants were called using samtools mpileup v1.9 (Li, et al. 2009) and ivar variant v1.2.2 (Grubaugh, et al. 2019), and consensus assemblies were generated using samtools mpileup v1.9 (Li, et al. 2009) and ivar consensus v1.2.2 (Grubaugh, et al. 2019). Only variants as the major allele were included in the final consensus assembly, and bases <10x were incorporated as N. Read mapping metrics were generated using samtools coverage v1.10 (Li, et al. 2009). Only samples with ≥80% genome coverage at ≥100x mean read depth were included in this study. B.1.1.7 lineages were determined using Pangolin v2.1.8 (https://github.com/cov-lineages/pangolin).

SARS-CoV-2 consensus assemblies for clinical samples with N501Y and/or S69-70del mutations are available on NCBI Genbank (MW452527-28, MW460886, MW494396-97, MW494398-99, MW545525, MW560187-88) and GISAID (EPI_ISL_790567-68, EPI_ISL_802724, EPI_ISL_852858-59, EPI_ISL_852860-61, EPI_ISL_906918, EPI_ISL_918493, EPI_ISL_918495).

## Data Availability

Relevent data are contained within the manuscript and SARS-CoV-2 consensus assemblies for clinical samples with N501Y and/or S69-70del mutations are available on NCBI Genbank, with accession IDs listed in the method section of the paper.

